# Physician attitudes about abortion at a Midwestern academic medical center

**DOI:** 10.1101/2020.05.08.20094540

**Authors:** Nicholas B. Schmuhl, Laurel W. Rice, Cynthia K. Wautlet, Jenny A. Higgins

**Author notes:** **Conflicts of interests:** The authors report no conflict of interest. **Funding source:** University of Wisconsin-Madison Office of the Vice Chancellor for Research and Graduate Education ($31,768). **Role of the funding source:** The funding source had no role in study design; in the collection, analysis and interpretation of data; in the writing of the report; or in the decision to submit the article for publication. **Paper Presentation:** These findings have been submitted to the Society of Family Planning 2020 Annual Meeting in Baltimore, Maryland, October 10 – 12, 2020. **Corresponding Author:** Laurel W. Rice, MD, University of Wisconsin-Madison, School of Medicine and Public Health, McConnell Hall, 4th Floor, 1010 Mound Street, Madison, WI 53715.

## Abstract

**Background:** Almost every medical professional organization supports abortion access. Meanwhile, federal and state-level policies continue to erode abortion-related healthcare. Physicians are instrumental to abortion access, and their evidence-based attitudes could significantly influence public understanding. However, most studies of physician attitudes about abortion focus on specific subgroups. A study of abortion attitudes among a broader population of clinicians is important for at least three reasons. First, results could provide insights and strategies to improve access and reduce stigma at academic medical centers and beyond. Second, findings could explain discrepancies between expressions of support for abortion by the medical community and the inability or unwillingness of the same community to provide sufficient access to abortion services. Third, gauging the climate of opinion among physicians in a politically contested state is likely to be informative given that most abortion-related judicial decisions will occur in state-versus federal-level courts, and physician attitudes could potentially influence public policy.

**Objective:** To use leading survey methodologies to assess abortion-related attitudes among all physicians at the largest academic medical center in a politically contested Midwestern state.

**Study Design:** Investigators developed a cross-sectional survey to gauge abortion-related knowledge, attitudes, and practices. The university’s survey research center disseminated the survey to all 1,357 physician faculty members of the school of medicine and public health using a web and mail mixed-mode methodology (67% response rate). Analyses included chi-squared tests and binary logistic regression models of support for abortion procedures and willingness to consult in abortion care.

**Results:** Across more than 20 specialties and all sociodemographic categories, physicians reported strong support for abortion. Majorities expressed support for medication (81%) and surgical abortion (80%), that abortion should be legal in all or most cases (88%), and that a state law banning abortion would make women’s health worse (91%). While nearly all physicians (94%) care for women of reproductive age, most (69%) reported no opportunity to participate in abortion care and fewer than half (44%) knew whom to contact to refer a patient for abortion care. Female physicians and those who considered their expertise relevant to abortion were more supportive, while physicians of color and highly religious physicians were less supportive. Few physicians reported participating in any aspect of abortion care (14%), though nearly two-thirds were willing to consult in such care (65%). Those with relevant expertise were more willing to consult, while physicians of color and highly religious physicians were less willing. While most physicians said they support unrestricted access to abortion (63%) and the efforts of abortion providers (70%) “a lot,” a majority perceived relatively less support among their professional peers, revealing a climate of pluralistic ignorance.

**Conclusions:** Despite overwhelming support for abortion among this population, participation in any aspect of abortion care is remarkably low. Physicians across all disciplines need clear training and guidelines on how to refer patients for abortion care, and abortion should be normalized and integrated into mainstream medicine. Given professional organizations’ support of abortion and physicians’ cultural influence, these results can be used to inform public policy regarding abortion access.

## Introduction

Forty-two percent of unintended pregnancies end in abortion,^1^ which is safe^2,3^ and has no known negative impacts on women’s physical or mental health beyond those of any surgical procedure.^4^ Access to abortion is associated with improved socioeconomic outcomes and relationship quality for women, as well as benefits to their current children.^5–8^ Almost every medical professional organization supports abortion access.^9–11^ However, federal and state-level policies continue to erode abortion-related healthcare^12,13^—including in Wisconsin.^14,15^ Indeed, without protections enacted after *Roe v. Wade*, Wisconsin’s pre-Civil War (1849) statute, which criminalizes abortion without exceptions for rape or incest, would become enforceable.^16^

Physicians are instrumental to abortion care and access.^12^ Doctors are more trusted cultural figures than politicians, lawyers, or even clergy members,^17^ and their evidence-based attitudes about abortion could significantly influence public understandings and policy. However, most studies of physician attitudes about abortion focus only on specific subgroups. Research suggests that most women’s health specialists^18^ and primary care providers^19^ support abortion access, though far fewer participate in any aspect of abortion care^20–22^—including referrals.^23,24^ Abortion providers frequently report feeling stigmatized at work,^25–27^ potentially contributing to the decline in access. Policies tend to protect physicians who conscientiously object to abortion,^28^ while rarely addressing the role of conscience in abortion provision.^29^

A study of abortion attitudes among a broader population of clinicians is important for at least three reasons. First, results could provide insights and strategies to improve access and reduce stigma at academic medical centers and beyond. Second, findings could help explain discrepancies between expressions of support for abortion by the medical community and the inability or unwillingness of the same community to provide sufficient access. Third and finally, gauging the climate of opinion among physicians in a politically contested state is likely to be informative given that most abortion-related judicial decisions will occur in state-versus federal-level courts.

## Materials and Methods

Investigators developed a cross-sectional survey of 45 questions gauging physicians’ knowledge, attitudes, and practices regarding abortion. Several of these items were replicated^26,30^ or adapted^26^ from previous studies. The intent was to field the survey to every currently-practicing physician faculty member at the University of Wisconsin-Madison School of Medicine and Public Health (UW SMPH), the largest medical school in the state of Wisconsin.

The UW SMPH Human Resources Department provided an up-to-date list of 1,357 practicing physician faculty members and their contact information, by permission of the Dean. The University of Wisconsin Survey Center (UWSC) consulted on survey methodology, assisted with formatting and production, and disseminated the survey using evidence-based web/mail mixed-mode methodology.^31^ First, they mailed individualized introductory letters outlining the purpose of the study containing $5 cash incentives and a unique URL and passcode for each participant to access the online questionnaire. One week later, those who had not responded received an email containing their unique URL and passcode, followed by up to three weekly reminder emails to non-responders. Finally, a paper questionnaire was mailed along with a stamped, self-addressed return envelope to those who had not responded after six weeks. The UWSC collected responses over three months in early 2019 and delivered a de-identified dataset to the study team.

An initial filter question asked participants to confirm that they were currently practicing faculty physicians at UW SMPH. Others were directed to the sociodemographics section at the end of the survey and excluded from all phases of analysis.

The University of Wisconsin-Madison Health Science Minimal Risk Institutional Review Board (IRB) deemed this study exempt from full review because the research is not federally supported, does not fall under Veterans Affairs regulations, and is not Food and Drug Administration-regulated. Nonetheless, the study underwent minimal IRB review to ensure adequate provisions were in place to protect privacy and maintain confidentiality.

### Measures

In order to facilitate statistical analyses and/or simplify interpretation of results, we combined or collapsed responses to several survey items into composite measures. The original response formats and their combined categories appear in Tables 1 through 3. Here, we present an overview of our reasoning behind some groupings and provide information about our primary attitude variables.

**Table 1:** Sociodemographic and Professional Characteristics of Faculty Physicians at the University of Wisconsin School of Medicine and Public Health, 2019 (N=893)

|  | N | % |
| --- | --- | --- |
| **Gender** |  |  |
| Female | 398 | 45.4 |
| Male | 476 | 54.3 |
| Other | 2 | 0.2 |
| **Age** |  |  |
| ≤39 | 235 | 27.3 |
| 40 - 49 | 288 | 33.5 |
| 50 - 59 | 196 | 22.8 |
| 60 - 69 | 123 | 14.3 |
| ≥70 | 19 | 2.2 |
| **Race and/or ethnicity** |  |  |
| Arab or Middle Eastern | 8 | 0.9 |
| Asian | 91 | 10.5 |
| Black or African American | 11 | 1.3 |
| Hispanic or Latino/a | 18 | 2.1 |
| Native American or Alaskan Native | 3 | 0.3 |
| Pacific Islander | 2 | 0.2 |
| White | 723 | 83.2 |
| Mixed or Other | 13 | 1.5 |
| **Parenting status** |  |  |
| Does not have children | 126 | 14.2 |
| Has children or participant/partner currently pregnant | 759 | 85.8 |
| **How much do you incorporate your religious beliefs into all your dealings in life?** |  |  |
| No religious identification | 425 | 48.3 |
| A little or not at all | 154 | 17.5 |
| Somewhat | 127 | 14.4 |
| Quite a bit or a great deal | 174 | 19.8 |
| **How often do you attend a church, temple, mosque, or other religious meetings?** |  |  |
| No religious identification | 425 | 48.4 |
| Once a year or less | 98 | 10.9 |
| At least a few times a year | 249 | 28.3 |
| Once a week or more | 109 | 12.4 |
| **Did you get any exposure to abortion care during your medical education, such as in medical school, residency or fellowship, even if you did not participate?** |  |  |
| Yes | 494 | 55.5 |
| No | 396 | 44.5 |
| **Do you see women of reproductive age in your practice?** |  |  |
| Yes | 838 | 94.3 |
| No | 51 | 5.7 |
| **Medical specialty** |  |  |
| Internal Medicine | 233 | 26.4 |
| Pediatrics | 118 | 13.3 |
| Family Medicine | 111 | 12.6 |
| Surgery | 99 | 11.2 |
| Radiology | 61 | 6.9 |
| Anesthesiology | 56 | 6.3 |
| Obstetrics and Gynecology | 39 | 4.4 |
| Emergency Medicine | 32 | 3.6 |
| Pathology | 27 | 3.1 |
| Neurology | 24 | 2.7 |
| Psychiatry | 19 | 2.1 |
| Dermatology | 15 | 1.7 |
| Ophthalmology | 13 | 1.5 |
| Radiation Oncology | 7 | 0.8 |
| Rehabilitation Medicine | 7 | 0.8 |
| Geriatrics | 6 | 0.7 |
| Urology | 5 | 0.6 |
| Genetics | 4 | 0.5 |
| Hospital Medicine | 4 | 0.5 |
| Medical Oncology | 2 | 0.2 |
| Otolaryngology | 2 | 0.2 |

**Table 2:**
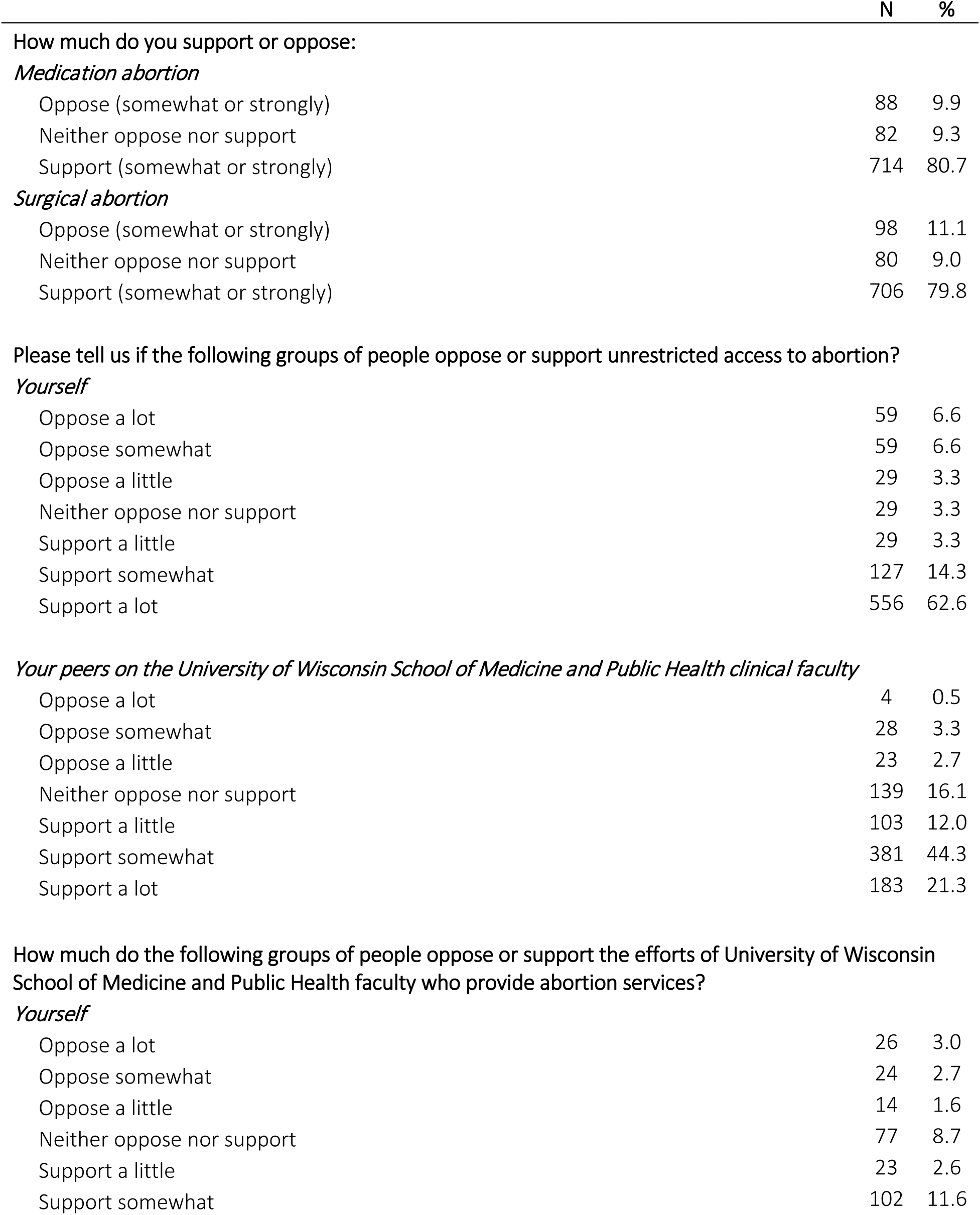
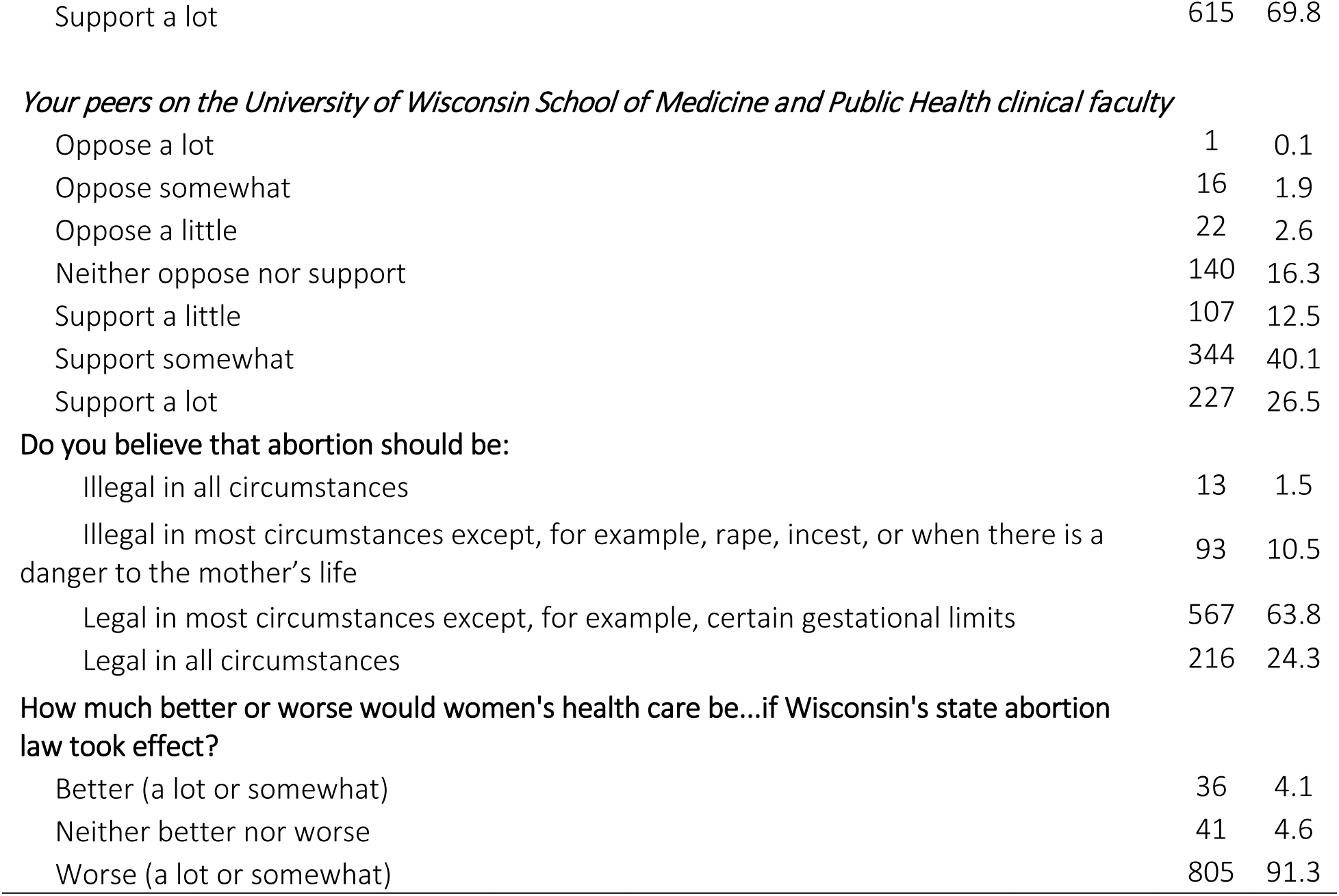
Attitudes and perceptions about abortion care, access, providers, and policy among faculty physicians at the University of Wisconsin School of Medicine and Public Health, 2019 (N=893)

|  | N | % |
| --- | --- | --- |
| **How much do you support or oppose:** |  |  |
| ***Medication abortion*** |  |  |
| Oppose (somewhat or strongly) | 88 | 9.9 |
| Neither oppose nor support | 82 | 9.3 |
| Support (somewhat or strongly) | 714 | 80.7 |
| ***Surgical abortion*** |  |  |
| Oppose (somewhat or strongly) | 98 | 11.1 |
| Neither oppose nor support | 80 | 9.0 |
| Support (somewhat or strongly) | 706 | 79.8 |
| **Please tell us if the following groups of people oppose or support unrestricted access to abortion?** |  |  |
| ***Yourself*** |  |  |
| Oppose a lot | 59 | 6.6 |
| Oppose somewhat | 59 | 6.6 |
| Oppose a little | 29 | 3.3 |
| Neither oppose nor support | 29 | 3.3 |
| Support a little | 29 | 3.3 |
| Support somewhat | 127 | 14.3 |
| Support a lot | 556 | 62.6 |
| ***Your peers on the University of Wisconsin School of Medicine and Public Health clinical faculty*** |  |  |
| Oppose a lot | 4 | 0.5 |
| Oppose somewhat | 28 | 3.3 |
| Oppose a little | 23 | 2.7 |
| Neither oppose nor support | 139 | 16.1 |
| Support a little | 103 | 12.0 |
| Support somewhat | 381 | 44.3 |
| Support a lot | 183 | 21.3 |
| **How much do the following groups of people oppose or support the efforts of University of Wisconsin School of Medicine and Public Health faculty who provide abortion services?** |  |  |
| ***Yourself*** |  |  |
| Oppose a lot | 26 | 3.0 |
| Oppose somewhat | 24 | 2.7 |
| Oppose a little | 14 | 1.6 |
| Neither oppose nor support | 77 | 8.7 |
| Support a little | 23 | 2.6 |
| Support somewhat | 102 | 11.6 |
| Support a lot | 615 | 69.8 |
| *Your peers on the University of Wisconsin School of Medicine and Public Health clinical faculty* |  |  |
| Oppose a lot | 1 | 0.1 |
| Oppose somewhat | 16 | 1.9 |
| Oppose a little | 22 | 2.6 |
| Neither oppose nor support | 140 | 16.3 |
| Support a little | 107 | 12.5 |
| Support somewhat | 344 | 40.1 |
| Support a lot | 227 | 26.5 |
| **Do you believe that abortion should be:** |  |  |
| Illegal in all circumstances | 13 | 1.5 |
| Illegal in most circumstances except, for example, rape, incest, or when there is a danger to the mother's life | 93 | 10.5 |
| Legal in most circumstances except, for example, certain gestational limits | 567 | 63.8 |
| Legal in all circumstances | 216 | 24.3 |
| **How much better or worse would women's health care be...if Wisconsin's state abortion law took effect?** |  |  |
| Better (a lot or somewhat) | 36 | 4.1 |
| Neither better nor worse | 41 | 4.6 |
| Worse (a lot or somewhat) | 805 | 91.3 |

**Table 3:** Participation in abortion-related care, consultancy, and advocacy among faculty physicians at the University of Wisconsin School of Medicine and Public Health, 2019 (N=893)

|  | N | % |
| --- | --- | --- |
| In your current role, do you participate in abortion care in any way? |  |  |
| Yes | 119 | 14.1 |
| No - no opportunity | 578 | 68.7 |
| No - choose not to participate | 144 | 17.1 |
| How relevant is your medical expertise to the issue of abortion? |  |  |
| Not at all | 252 | 28.4 |
| A little or somewhat | 454 | 51.1 |
| Very or extremely | 182 | 20.5 |
| How willing would you be to be a consultant in the care of any patient seeking abortion? |  |  |
| Not at all | 76 | 8.7 |
| A little or somewhat | 231 | 26.4 |
| Very or extremely | 568 | 64.9 |
| If you had to refer a patient for abortion care, would you know whom to contact? |  |  |
| Yes | 367 | 43.9 |
| No | 469 | 56.1 |
| How concerned are you that if you take a strong public stance either in support of or against abortion you will: |  |  |
| *Alienate some patients* |  |  |
| Not at all | 175 | 21.4 |
| A little or somewhat | 487 | 59.5 |
| Very or extremely | 156 | 19.1 |
| *Alienate some co-workers* |  |  |
| Not at all | 208 | 25.2 |
| A little or somewhat | 488 | 59 |
| Very or extremely | 131 | 15.8 |
| *Be harassed or harmed by protestors of opposing views* |  |  |
| Not at all | 138 | 17.4 |
| A little or somewhat | 488 | 61.7 |
| Very or extremely | 165 | 20.9 |

Physicians were asked to choose their area of practice from 13 predetermined choices, or “Something else – please tell us,” which included a space for text entry. Of the 20 specialties listed or typed in, we created three collapsed categories to account for differences in training, practices, patient populations, and exposure to abortion across disciplines while protecting the confidentiality of physicians who practice less common specialties. These three practice categories included “office-based” practices (internal medicine, geriatrics, pediatrics, family medicine, neurology, psychiatry, dermatology, ophthalmology, speech pathology, rehabilitation medicine, palliative care, and medical oncology; 62%), “surgical” practices (surgery, obstetrics and gynecology, urology, and otolaryngology; 16%), and “other” practices (emergency medicine, radiology, anesthesiology, pathology, genetics, radiation oncology, and hospital medicine; 22%).

Separate items gauged participants’ support for medication abortion and surgical abortion on five-point scales ranging from “strongly support” to “strongly oppose,” with “in between” as a midpoint. Levels of support for both abortion procedures were highly positively correlated, *r*(882)=0.96, p<0.001. For the purposes of further analyses, the variables were combined to create a binary variable consisting of those who registered any opposition or neutrality (no support, 20%), and those who “somewhat” or “strongly support” abortion (any support, 80%).

A single item queried participants about the perceived relevance of their medical expertise to the issue of abortion on a five-point Likert-type scale, ranging from “not at all” to “extremely.” These responses were collapsed in order to create a binary variable consisting of those who felt that their expertise was “not at all” to “somewhat” relevant (low relevance, 80%) and those who felt that their expertise was “very” or “extremely” relevant (high relevance, 20%).

The questionnaire measured willingness to consult in the care of any patient seeking abortion via a single item with a five-point Likert-type scale ranging from “not at all” to “extremely.” We dichotomized responses into a binary variable consisting of those who were “not at all” to “somewhat” willing (low willingness, 35%) or “very” or “extremely” willing (high willingness, 65%).

The survey queried participants about their own support for “unrestricted access to abortion” and “the efforts of UW SMPH faculty who provide abortion services,” and also asked them to estimate support for the same concepts among “your peers on the UW SMPH clinical faculty.” The seven-point Likert-type response scales for these items ranged from “oppose a lot” to “support a lot,” with “neither oppose nor support” as a midpoint. We created perceived self-peer difference scores for these items by subtracting estimated peer support from participants’ self-reported support. Those with negative difference scores perceived themselves as less supportive than professional peers. Those with positive difference scores perceived themselves as more supportive than professional peers. The remainder perceived no difference in support.

Three survey items focused on participants’ religiosity, including measures of religious identification, religious influence on daily life, and attendance at religious services or meetings. We combined these items into a binary religiosity variable: 1) those who indicated no religious identification or religious identifiers who fell below the midpoint of the combined religiosity scale (low religiosity; 74%); and 2) religious identifiers who fell in the upper half of the scale (high religiosity; 26%).

Participants reported whether they currently had children or other dependents, and whether they or a partner was currently pregnant.

### Analyses

Investigators used IBM SPSS Statistics 26 to perform analyses. At a bivariate level, when comparing responses to various survey items by sociodemographic or professional characteristics, we used chi-square tests of independence to confirm that observed differences were statistically significant.

At the multivariate level, we used binary logistic regression to model two outcomes of interest: 1) support for abortion care procedures; and 2) willingness to consult in abortion care. Socio-demographic control variables included age, gender, race/ethnicity, and parenting status. Covariates, selected based on both previous literature^26^ and significant bivariate chi-square associations, included exposure to abortion during training and perceived relevance of medical expertise to the issue of abortion. Because interdisciplinarity is a unique feature of the study sample, we stratified models by practice type in order to elucidate potential differences between physicians of different backgrounds and expertise.

## Results

Nine hundred and thirteen of 1,357 distributed surveys were returned, for an adjusted response rate of 67%. Nearly all participants (N=893; 98%) confirmed that they were currently practicing physicians; others were excluded from further analysis.

### Socio-demographic and professional characteristics

As shown on Table 1, a slight majority of physicians identified as male (54%). Respondents tended to be middle-aged (M=47.7 years; SD=10.7) and white (83%). Most had children and/or reported that they or a partner were currently pregnant (86%). Slightly more than half (52%) said they identified with a religion, and religious identifiers tended to report a moderate influence of religious beliefs on their daily lives and occasional attendance at religious meetings.

Participants represented more than 20 medical specialties. Nearly all (94%) said they see women of reproductive age in their practices. Over half (56%) reported some exposure to abortion at any point during their medical training.

### Attitudes and perceptions about abortion and abortion providers

Table 2 includes responses to survey items regarding attitudes about abortion care, access, providers, and policy; Figure 1 visually illustrates a subset of items contained in Table 2. As indicated in both, we found strong support for abortion among UW SMPH faculty physicians across various measures, ranging from 80% who support surgical abortion to 91% who said that women’s health care in Wisconsin would be worse if *Roe v. Wade* were overturned and the state law took effect (91%).

**Figure 1:**
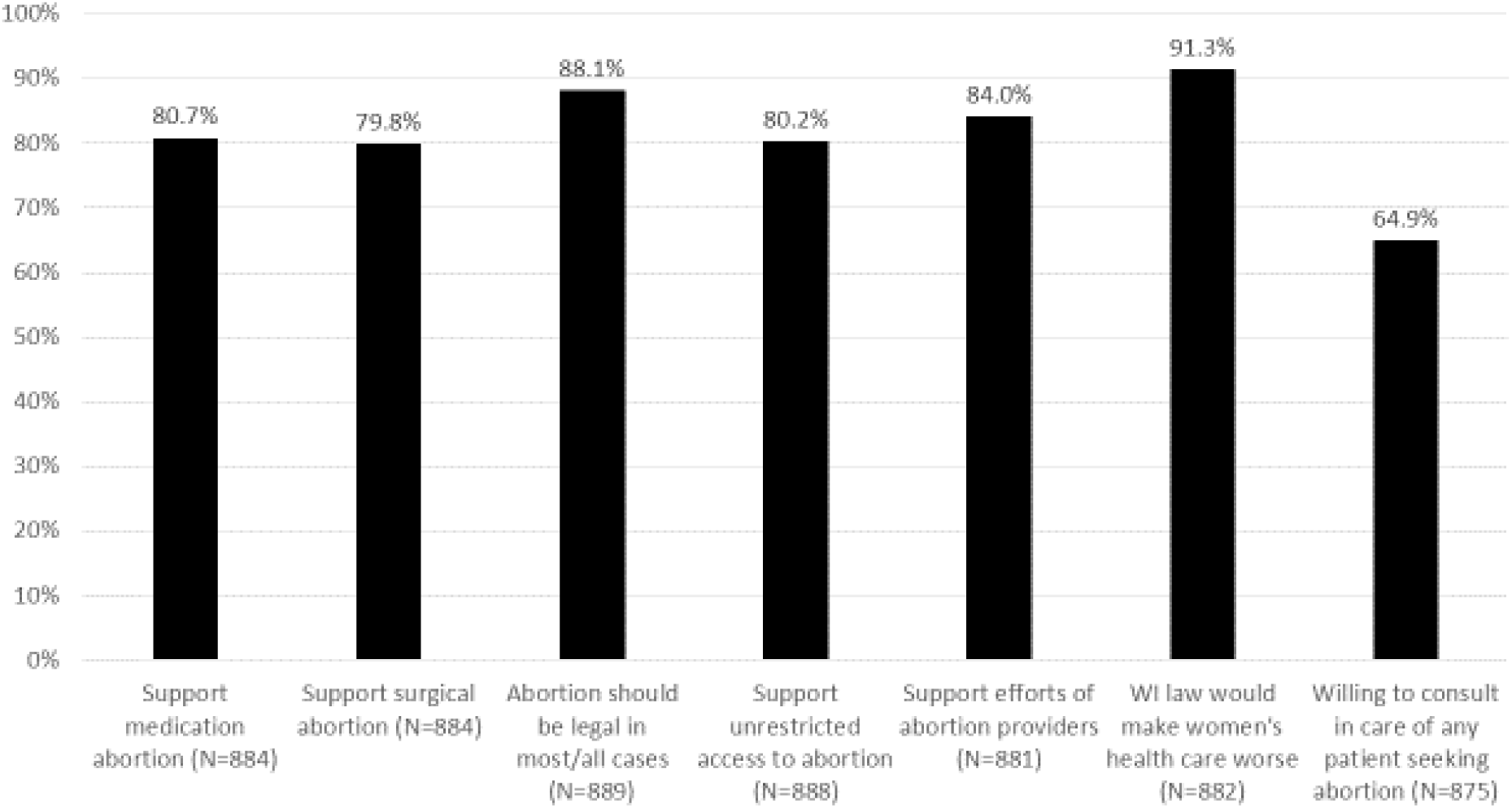
Multiple measures of physician support for abortion; University of Wisconsin School of Medicine and Public Health, 2019

In terms of participants’ self-reported support for unrestricted access to abortion and the efforts of UW SMPH faculty who provide abortions, majorities placed their own attitudes on the supportive end of the response spectrum for both unrestricted access to abortion (80%) and the efforts of abortion providers (84%). However, they underestimated support among their professional peers. While most participants said they support access (63%) and providers (70%) “a lot,” they most commonly estimated that their peers only “somewhat” supported access (44%) and providers (40%). Notably, while few participants claimed neutrality (“neither oppose nor support”) regarding access (3%) or providers (9%), they perceived greater neutrality among their peers (16% for both access and providers). Chi-square tests of independence revealed statistically significant differences between self-appraisals and perceptions of peer support for abortion access (*X*^2^ (36, N=861) = 363.19, p<0.001) and the efforts of abortion providers (*X*^2^ (36, N=861) = 363.19, p<0.001).

Figure 2 presents comparisons (i.e., difference scores) between self-reported attitudes about abortion and estimates of peers’ attitudes. More than half of physicians in our sample perceived themselves as more supportive of abortion than their professional peers (51% for both access and providers), indicating a split between actual climate of opinion and perceived climate of opinion (i.e., pluralistic ignorance).

**Figure 2:**
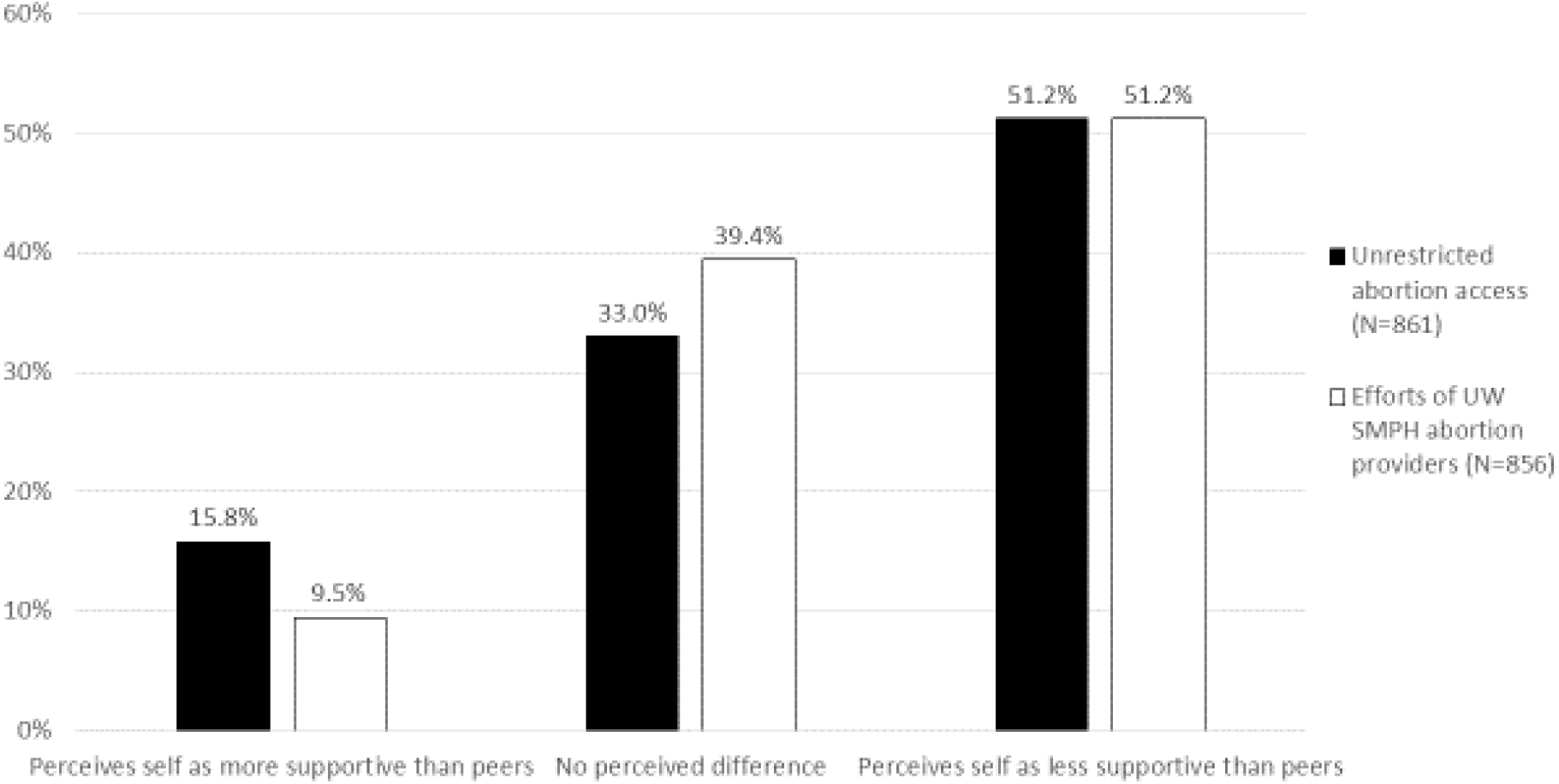
Physician abortion attitudes compared to perceived attitudes of professional peers; University of Wisconsin School of Medicine and Public Health, 2019

### Involvement in abortion care

As indicated in Table 3, few physicians said they participate in abortion care in any way in their current role (14%), though a majority (72%) reported that their medical expertise was relevant to abortion to some extent, and nearly two thirds said they were either “very” or “extremely” willing to consult in the care of any patient seeking abortion (65%). Meanwhile, fewer than half said that they would know whom to contact if they had to refer a patient for abortion care (44%).

Majorities were at least “a little” concerned that if they were to take a public stand either for or against abortion, they would alienate some patients (79%), alienate some co-workers (75%), or be harassed or harmed by protestors of opposing views (83%).

### Factors associated with support for abortion

Table 4 presents results from binary logistic regression models with support for abortion as the outcome. Female physicians were more likely than male physicians to be supportive of abortion in the overall population (OR 1.87, 95% CI 1.25-2.80) and among those with surgical (OR 6.07, 95% CI 1.46-25.18) practices. Physicians identifying as persons of color or mixed race were significantly less likely than those identifying as white to support abortion, both in the overall population (OR 0.51, 95% CI 0.32-0.81) and among those with office-based practices (OR 0.43, 95% CI 0.24-0.76). Highly-religious physicians were also significantly less likely to than non- or less-religious physicians to support abortion, both in the overall population (OR 0.16, 95% CI 0.10-0.24) and among those with office-based (OR 0.20, 95% CI 0.12-0.34), surgical (OR 0.11, 95% CI 0.03-0.35), and other types of practices (OR 0.01, 95% CI 0.04-0.25). Those who considered their expertise moderately-to-highly relevant to the issue of abortion were significantly more likely than their counterparts to support abortion in the overall population (OR 2.10, 95% CI 1.23-3.61), as well as those with office-based practices (OR 2.02, 95% CI 1.04-3.91).

**Table 4.**
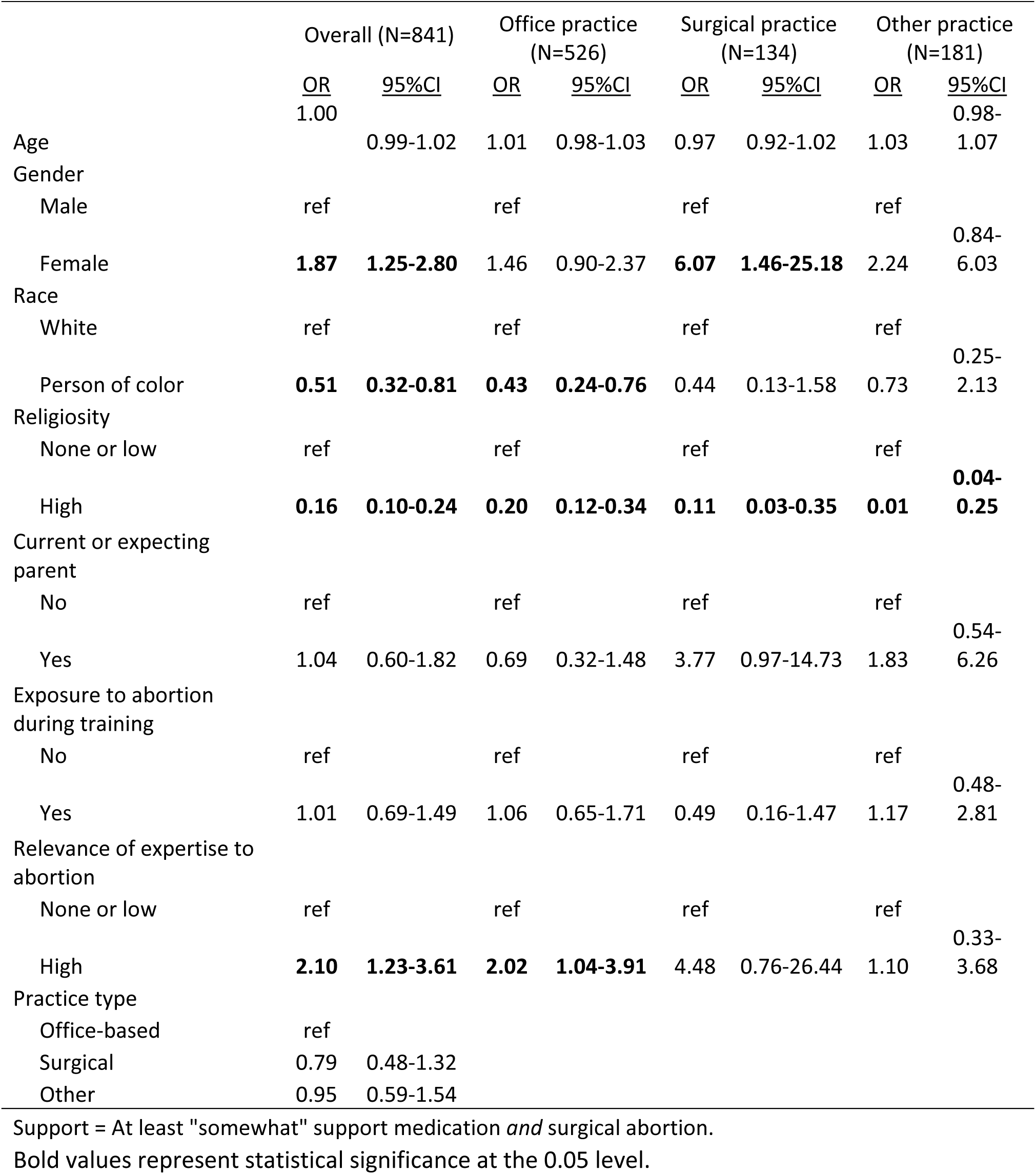
Factors associated with support for medical and surgical abortion among faculty physicians, University of Wisconsin School of Medicine and Public Health, 2019 (N=841)

### Factors associated with willingness to consult in abortion care

Table 5 presents results from multivariate logistic regression models with willingness to consult in abortion care as the outcome. In general, physicians with practices categorized as “other” were significantly less likely to be willing to consult in abortion care than those with office-based practices (OR 0.64, 95% CI 0.44-0.93). The difference between surgical and office-based practices was not statistically significant.

**Table 5.** Factors associated with willingness to consult in the care of any patient seeking abortion, faculty physicians at the University of Wisconsin School of Medicine and Public Health, 2019 (N=835)

|  | Overall (N=835) |  | Office practice (N=523) |  | Surgical practice (N=133) |  | Other practice (N=179) |  |
| --- | --- | --- | --- | --- | --- | --- | --- | --- |
|  | <u>OR</u> | <u>95%CI</u> | <u>OR</u> | <u>95%CI</u> | <u>OR</u> | <u>95%CI</u> | <u>OR</u> | <u>95%CI</u> |
| Age | **0.98** | **0.97-0.998** | **0.97** | **0.95-0.99** | 1.01 | 0.97-1.05 | 1.00 | 0.97-1.03 |
| Gender |  |  |  |  |  |  |  |  |
| Male | ref |  | ref |  | ref |  | ref |  |
| Female | 0.87 | 0.63-1.20 | 0.72 | 0.48-1.08 | 2.54 | 0.99-6.54 | 0.74 | 0.36-1.55 |
| Race |  |  |  |  |  |  |  |  |
| White | ref |  | ref |  | ref |  | ref |  |
| Person of color | **0.48** | **0.32-0.71** | **0.50** | **0.30-0.83** | **0.33** | **0.11-0.96** | **0.40** | **0.17-0.97** |
| Religiosity |  |  |  |  |  |  |  |  |
| None or low | ref |  | ref |  | ref |  | ref |  |
| High | **0.40** | **0.29-0.54** | **0.54** | **0.36-0.80** | **0.40** | **0.18-0.91** | **0.15** | **0.07-0.32** |
| Parent or pregnant |  |  |  |  |  |  |  |  |
| No | ref |  | ref |  | ref |  | ref |  |
| Yes | 1.04 | 0.66-1.64 | 0.88 | 0.48-1.60 | 1.82 | 0.65-5.08 | 1.02 | 0.36-2.85 |
| Exposure to abortion during training |  |  |  |  |  |  |  |  |
| No | ref |  | ref |  | ref |  | ref |  |
| Yes | 1.25 | 0.91-1.70 | 1.15 | 0.77-1.71 | 1.27 | 0.54-2.98 | 1.66 | 0.82-3.36 |
| Relevance of expertise to abortion |  |  |  |  |  |  |  |  |
| None or low | ref |  | ref |  | ref |  | ref |  |
| High | **5.26** | **3.18-8.70** | **5.89** | **3.08-11.29** | **3.44** | **1.06-11.15** | **7.18** | **2.04-25.31** |
| Practice type |  |  |  |  |  |  |  |  |
| Office-based | ref |  | ref |  | ref |  | ref |  |
| Surgical | 0.76 | 0.49-1.17 |  |  |  |  |  |  |
| Other | **0.64** | **0.44-0.93** |  |  |  |  |  |  |

Willingness to consult = "Very" or "extremely" willing to consult.
Bold values represent statistical significance at the 0.05 level.

Compared to white physicians, physicians who identified as persons of color were less likely to be willing to consult in abortion care in the overall population (OR 0.40, 95% CI 0.29-0.54), and also among those with office-based (OR 0.54, 95% CI 0.36-0.80), surgical (OR 0.40, 95% CI 0.18-0.91), and other types of practices (OR 0.40, 95% CI 0.17-0.97). More religious physicians were less willing to consult in abortion care in the overall population (OR 0.40, 95% CI 0.29-0.54), and among those with office-based (OR 0.54, 95% CI 0.36-0.78), surgical (OR 0.40, 95% CI 0.18-0.91), and other types of practices (OR 0.15, 95% CI 0.07-0.32). Physicians who felt their medical expertise was relevant to the issue of abortion were more likely to be willing to consult in abortion care in the overall population (OR 5.26, 95% CI 3.18-8.70), and among those with office-based (OR 5.89, 95% CI 3.08-11.29), surgical (OR 3.44, 95% CI 1.06-11.15), and other types of practices (OR 7.18, 95% CI 2.04-25.31).

## Comment

### Principle Findings

Across all sociodemographic factors and types of medical practices, physicians in this population overwhelmingly support abortion care, abortion providers, and pro-choice policies. Notably, the most commonly shared sentiment was that women’s health care would be worse in Wisconsin if the *Roe v. Wade* decision gave way to the restrictive state law that has remained unchanged for 170 years (91%).

### Results

Despite predominant support across all subgroups, some physicians were more likely to support abortion access than others. Women were generally more likely to support abortion than men—particularly women with surgical practices, who were six times as likely to support abortion as their male colleagues with surgical practices. Highly religious physicians were less likely to support abortion, as might be expected in any population. Physicians of color, who were underrepresented in this sample, were less likely than white physicians to support abortion, which may reflect concerns about reproductive autonomy grounded in a history of reproductive coercion among populations of color in the United States.

Given the level of stated support for abortion, direct participation in any aspect of abortion care is remarkably low (14%). While it is encouraging that nearly two-thirds (65%) are willing to consult in abortion cases, roughly the same proportion perceive that they have no opportunity to participate in abortion care (69%). The fact that fewer than half of physicians would know whom to contact if they had to refer a patient for abortion care (44%) is difficult to reconcile—especially given that almost every physician in our sample said they care for women of reproductive age (94%).

### Implications

We argue as others have that physicians across all specialties need clear training and guidelines on how and where to refer patients for abortion care^32^—in direct opposition to the recent federal Title X “gag rule” that prohibits healthcare providers at funded clinics from counseling or referring clients for abortion care.^33^ We also maintain that abortion care must become fully integrated into mainstream medicine, particularly given the widespread ability of physicians from various specialties to learn to prescribe and manage medication abortion.

In a population highly supportive of abortion, and one in which abortion opponents acknowledge that they are in the minority, the gap between stated support and participation (or willingness to participate by referring or consulting according to one’s professional skillset) bears consideration. It is illuminating that many physicians in this population underestimate support for abortion and overestimate neutrality among their colleagues, suggesting pluralistic ignorance and stigma. It is well established that abortion providers feel stigmatized by their colleagues^25^. In a climate where perceptions undercut actual support for abortion, or where abortion care is isolated to the extent that many physicians assume their colleagues have no opinion at all, supportive physicians may be less inclined to convert their attitudes into actions.

### Strengths and Limitations

This study expands the existing literature regarding healthcare provider attitudes about abortion by gauging the climate of opinion among physicians in all academic specialties at the largest medical training center in a state where abortion rights are threatened. While our sample is limited to a single academic medical center, this study can and should be replicated at similar institutions across the country. Our unusually strong response rate for a physician survey suggests that physicians are willing—perhaps eager—to express their attitudes about abortion.

### Conclusions

Strategies to reduce pluralistic ignorance and stigma include challenging inaccurate perceptions, promoting dialogue, and normalizing the accurate beliefs and experiences of group members.^34^ To that end, a next step toward reducing pluralistic ignorance and stigma surrounding abortion at UW SMPH, which can be replicated at every academic medical center, is to collect and broadly disseminate accurate information about the climate of opinion. Training and other interventions may be required to activate physician support, so that every physician who cares for women of reproductive age feels prepared to assist or refer any patient seeking abortion.

## Data Availability

Please contact the study authors with inquiries about data.

## Acknowledgments

The authors would like to thank Lisa Harris, Lisa Martin, and Meghan Seewald at the University of Michigan for consulting on the content of our survey instrument. We are also grateful to the University of Wisconsin Survey Center for their invaluable methodological expertise and multiple reviews of the survey instrument. Finally, we would like to acknowledge early contributions to this work by Helen Zukin (literature review) and Kelsey Wright (data analysis) at the University of Wisconsin-Madison.

## References

1. Finer LB, Zolna MR. Declines in Unintended Pregnancy in the United States, 2008–2011. N Engl J Med. 2016;374(9):843–852.

2. National Academies of Sciences E. The Safety and Quality of Abortion Care in the United States. National Academies Press; 2018.

3. Raymond EG, Grimes DA. The Comparative Safety of Legal Induced Abortion and Childbirth in the United States. Obstet Gynecol. 2012;119(2):215.

4. Brown SS, Eisenberg L. The Best Intentions: Unintended Pregnancy and the Well-Being of Children and Families. National Academies Press; 1995.

5. Upadhyay UD, Aztlan-James EA, Rocca CH, Foster DG. Intended pregnancy after receiving vs. being denied a wanted abortion. Contraception. 2019;99(1):42–47.

6. Foster DG, Biggs MA, Ralph L, Gerdts C, Roberts S, Glymour MM. Socioeconomic Outcomes of Women Who Receive and Women Who Are Denied Wanted Abortions in the United States. Am J Public Health. 2018;108(3):407–413.

7. Foster DG, Biggs MA, Raifman S, Gipson J, Kimport K, Rocca CH. Comparison of Health, Development, Maternal Bonding, and Poverty Among Children Born After Denial of Abortion vs After Pregnancies Subsequent to an Abortion. JAMA Pediatr. 2018;172(11):1053–1060.

8. Foster DG, Raifman SE, Gipson JD, Rocca CH, Biggs MA. Effects of Carrying an Unwanted Pregnancy to Term on Women’s Existing Children. J Pediatr. 2019;205:183–189.e1.

9. American Academy of Family Physicians. Frontline Physicians Call on Politicians to End Political Interference in the Delivery of Evidence Based Medicine. aafp.org.

10. AMA lawsuit to protect patient-physician relationship in North Dakota. Am Med Assoc.

11. AMA opposes proposed rule on Title X family planning program. Am Med Assoc.

12. Nash E. Abortion Rights in Peril — What Clinicians Need to Know. N Engl J Med. 2019;381(6):497–499.

13. Jones RK, Jerman J. Abortion Incidence and Service Availability In the United States, 2014. Perspect Sex Reprod Health. 2017;49(1):17–27.

14. State Facts About Abortion Wisconsin. Guttmacher Institute; 2019: 2.

15. Bad Medicine: How a Political Agenda Is Undermining Abortion Care and Access (3rd Ed.). National Partnership for Women & Families; 2018.

16. Wis. Stats. § 940.04. Abortion.

17. Gallup, Inc. Nurses Again Outpace Other Professions for Honesty, Ethics. Gallup.com. December 2018.

18. Dodge LE, Haider S, Hacker MR. Attitudes toward Abortion among Providers of Reproductive Health Care. Womens Health Issues. 2016;26(5):511–516.

19. Chuang CH, Martenis ME, Parisi SM, et al. Contraception and abortion coverage: what do primary care physicians think? Contraception. 2012;86(2):153–156.

20. Desai S, Jones RK, Castle K. Estimating abortion provision and abortion referrals among United States obstetrician-gynecologists in private practice. Contraception. 2018;97(4):297–302.

21. Stulberg DB, Dude AM, Dahlquist I, Curlin FA. Abortion Provision Among Practicing Obstetrician–Gynecologists: Obstet Gynecol. 2011;118(3):609–614.

22. Myran DT, Carew CL, Tang J, Whyte H, Fisher WA. Medical Students’ Intentions to Seek Abortion Training and to Provide Abortion Services in Future Practice. J Obstet Gynaecol Can. 2015;37(3):236–244.

23. Homaifar N, Freedman L, French V. “She’s on her own”: a thematic analysis of clinicians’ comments on abortion referral. Contraception. 2017;95(5):470–476.

24. Holt K, Janiak E, McCormick MC, et al. Pregnancy Options Counseling and Abortion Referrals Among US Primary Care Physicians: Fam Med. 2017: 10.

25. Martin LA, Debbink M, Hassinger J, Youatt E, Harris LH. Abortion providers, stigma and professional quality of life. Contraception. 2014;90(6):581–587.

26. Martin LA, Seewald M, Johnson TRB, Harris LH. Trusted Colleagues or Incompetent Hacks? Development of the Attitudes About Abortion-Providing Physicians Scale. Womens Health Issues. October 2019.

27. Britton LE, Mercier RJ, Buchbinder M, Bryant AG. Abortion providers, professional identity, and restrictive laws: A qualitative study. Health Care Women Int. 2017;38(3):222–237.

28. Morrell KM, Chavkin W. Conscientious objection to abortion and reproductive healthcare: a review of recent literature and implications for adolescents. Curr Opin Obstet Gynecol. 2015;27(5):333.

29. Harris LH. Recognizing Conscience in Abortion Provision. N Engl J Med. 2012;367(11):981–983.

30. Harris LH, Cooper A, Rasinski KA, Curlin FA, Lyerly AD. Obstetrician–Gynecologists’ Objections to and Willingness to Help Patients Obtain an Abortion. Obstet Gynecol. 2011; 118(4): 905–912.

31. Dykema J, Jones NR, Piché T, Stevenson J. Surveying Clinicians by Web: Current Issues in Design and Administration. Eval Health Prof. 2013;36(3):352–381.

32. Zurek M, O’Donnell J, Hart R, Rogow D. Referral-making in the current landscape of abortion access. Contraception. 2015;91(1):1–5.

33. Office of the Assistant Secretary for Health, Office of the Secretary, U.S. Department of Health and Human Services (HHS). Compliance With Statutory Program Integrity Requirements. Fed Regist. 2019;84(42):7714–7791.

34. Karaffa KM, Koch JM. Stigma, Pluralistic Ignorance, and Attitudes Toward Seeking Mental Health Services Among Police Officers. Crim Justice Behav. 2016;43(6):759–777.

